# Effects of early childhood education on early childhood development in Bangladesh

**DOI:** 10.1101/2023.12.16.23300090

**Authors:** Shimlin Jahan Khanam, Md Badsha Alam, Md Nuruzzaman Khan

## Abstract

**Background:** Early childhood education is considered as vital for the early childhood development (ECD) instead of scarcity of the relevant literature. This study aims to investigate the relationship between early childhood education and the early childhood development index (ECDI) in Bangladesh.

**Methods:** We analyzed data from 9,420 children (aged 3 and 4) extracted from the Multiple Indicator Cluster Survey conducted in 2019. The outcome variable considered was the Early Childhood Development Index, a composite index generated based on the responses to 10 items and categorized as children either developmentally on track (coded as 1) or not (coded as 0). Four domains of ECDI—physical, learning, emotional, and social well-being— were also considered as outcome variable. Attendance of the early childhood education program was the major exposure variable. The association between explanatory and outcome variables was determined using a multilevel logistic regression model, adjusting for potential covariates.

**Results:** Only one in every five children in Bangladesh was found to be enrolled in early childhood education. Non-participation in early childhood education was associated with a 44% lower likelihood (aOR, 0.56, 95% CI, 0.47-0.66) of positive ECDI compared to participation in early childhood education. Additionally, non-attendance of early childhood education, compared to attendance, was associated with lower odds in literacy-numeracy (aOR, 0.25, 95% CI, 0.21-0.28) and learning (aOR, 0.59, 95% CI, 0.45-0.77) domains of ECDI.

**Conclusion:** The findings provide evidence of the importance of early childhood education programs in ensuring early childhood development. This indicates the necessity of strengthening early childhood education programs in Bangladesh to ensure the overall well-being of children.

## Background

Early childhood development (ECD) is a crucial phase that encompasses the critical years from birth to age eight, representing a period of rapid physical, cognitive, and socio-emotional growth [1]. The experiences and environment during these formative years have profound and lasting effects on a child’s overall well-being and future outcomes [2, 3]. Research consistently indicates that positive ECD significantly contributes to cognitive skills, language proficiency, and social competence, laying the foundation for academic success and emotional resilience [2-4]. However, regardless of the importance of ensuring ECD, over 250 million children worldwide fail to reach their developmental potential, with the majority residing in low- and middle-income countries (LMICs) [5]. The prevalence is even higher in Asian and African countries [5]. The underlying reasons are that parents in these regions are mostly unaware of the importance of care during the early years, as well as the lack of early childhood development programs, including early childhood education [6, 7]. Other factors associated with poor ECD are poor socio-demographic, environmental, and health-related factors [8-11].

Early childhood education is a foundational component crucial for the comprehensive development of children [12]. Several countries at the global level have found its effective role in improving ECD [11, 13, 14]. Consequently, the government of Bangladesh, in 2003, initiated a program to provide early childhood education through over 65,500 primary schools located across Bangladesh [15]. Several non-governmental organizations also started providing early childhood education through their broad network [15]. These efforts even more extended in 2015 when the Sustainable Development Goals were established with particular focus on ECD [16]. Other LMICs also had a similar focus. However, the extent to which early childhood education programs influence ECD is mostly unexplored, with few studies conducted in developed countries and LMICSs and none in Bangladesh [6, 13, 14]. The findings of these studies are also scattered with significant and lack of association [6, 13, 14]. The reasons for such variations include the analysis of small-scale data and consideration of inadequate and inappropriate lists of confounder factors [6, 14]. To address these limitations, we conducted this study to explore the association between early childhood education and ECD, adjusted for covariates.

## Methods

### Study design and sampling procedure

The data for this study was extracted from the Multiple Indicator Cluster Survey (MICS) conducted in 2014, a nationally representative survey conducted every four years as part of UNICEF’s initiative to collect data in low- and middle-income countries (LMICs). The Bangladesh Bureau of Statistics conducted this survey in Bangladesh as a local implementation partner. The survey utilized a two-stage stratified random sampling approach to select nationally representative households. In the first stage, a total of 3,220 primary sampling units (PSUs) were selected through random sampling from the 293,579 PSUs generated during the 2011 Bangladesh national population census. In the second stage, 20 households were systematically chosen from each PSU using probability proportional to the sampling size. This process resulted in a list of 64,400 households, of which the survey was conducted in 52,771 households with a response rate of 81.94%. Among the selected households, there were 24,686 eligible children meeting the following conditions: (i) children aged 5 years or below, and (ii) residing in the selected households. Data were ultimately collected from 23,402 children. Additional details about the sampling procedure have been published elsewhere [17].

### Analytical sample

We analysed a total of 9,420 data from children aged 3 to 4 for this study from the main survey. The basis for this selection was their eligibility for enrolment to collect the necessary data needed to generate the Early Childhood Development Index (ECDI), as well as the availability of information regarding early childhood education [18].

### Outcome variable

The primary outcome variable considered was ECDI-a compositive index generated based on the 10 items response and categorized as children either developmentally track (1) or not (0). Total of 10 items were considered to generate this index under the four domains: literacy-numeracy (three items), physical (two items), socio-emotional (three items), and learning (two items) [18]. These four domains were also considered as outcome variables. Details of these items are presented in Supplementary table 1. To be classified as developmentally on track in a specific domain, children needed to provide positive responses to at least two items in literacy-numeracy and socio-emotional domains, as well as at least one item in the physical and learning domains [18, 19]. These domain-specific classifications were then combined to generate the overall ECDI, where children who were developmentally on track in at least three out of the four domains were considered developmentally track. Detailed information regarding the generation of these indexes can be found elsewhere [18].

### Exposure variable

The primary exposure variable considered was attendance in the early childhood education program (yes, no). The relevant data were collected by asking parents or actual caregivers of the selected children about whether the index children were enrolled in early childhood education. It is important to mention that early childhood education has been a governmental priority issue in Bangladesh over the decades. The government, as well as the ongoing private sectors, are the main providers of this education.

### Covariates

We considered various factors to account for the relationship between ECDI and early childhood education. These factors were chosen after reviewing relevant literature in Bangladesh and other LMICs [2, 4, 6, 8-11]. They included the child’s age (3 years, 4 years), child’s gender (male, female), mother’s education level (pre-primary or none, primary, secondary, higher secondary, and more), and whether the mother faced any functional difficulties (yes, no). Household level factors included were household wealth quintile index. Community-level characteristics included the place of residence (urban, rural) and division of residence (Barishal, Chattogram, Dhaka, Khulna, Mymensingh, Rajshahi, Rangpur, Sylhet).

### Statistical analysis

We used descriptive statistics to outline the characteristics of the respondents. Multilevel logistic regression model was used to determine the association between exposure and outcome variable adjusted for covariates. The reasons for using multilevel modelling was nested structure of the data we analyzed where children were nested within households, and households were nested within clusters (PSUs). This clustering creates interdependence among the data, making the use of a simple logistic regression model methodologically less accurate and potentially leading to underestimation or overestimation of the true associations [24]. Separate models were applied for the ECDI and its four domains. The results are presented as Odds Ratios (OR) with their corresponding 95% Confidence Intervals (CI). All statistical analyses were carried out using Stata version 15.1.

## Results

### Background characteristics of the respondents

The demographic characteristics of the respondents are presented in Table 1. Participation in the early childhood education program was reported by 19.3% of the total children analyzed. The distribution of children’s age was relatively equal, with 51.1% at 3 years and 48.9% at 4 years. Approximately 51.8% were male, and 48.2% were female. Mother’s education levels exhibited variation, with 13.2% having pre-primary education, 24.3% with primary education, 48.1% with secondary education, and 14.4% with higher education. About one-quarter of the total children resided in the poorest wealth quintile. Nearly 21% of the total children were from urban areas. The regional distribution revealed varying percentages across different areas, with Dhaka reporting the highest percentage (23.1%), while Sylhet reported the lowest (8.0%).

**Table 1:** Background characteristics of the respondents, MICS, 2019 (N= 9,420).

| **Demographics of children** | **%** | **95% CI** |
| --- | --- | --- |
| **Attended in early childhood education** |  |  |
| Yes | 19.3 | 18.4-20.3 |
| No | 80.7 | 79.7-81.6 |
| **Child's age** |  |  |
| 3 years | 51.1 | 50.0-52.2 |
| 4 years | 48.9 | 47.8-50.0 |
| **Child's gender** |  |  |
| Male | 51.8 | 50.6-52.9 |
| Female | 48.2 | 47.1-49.4 |
| **Mother's education** |  |  |
| Pre-primary | 13.2 | 12.4-14.1 |
| Primary | 24.3 | 23.3-25.4 |
| Secondary | 48.1 | 46.9-49.3 |
| Higher | 14.4 | 13.6-15.3 |
| **Wealth quintile** |  |  |
| Poorest | 22.4 | 21.2-23.6 |
| Second | 19.9 | 19.0-20.9 |
| Middle | 18.6 | 17.7-19.6 |
| Fourth | 19.3 | 18.3-20.4 |
| Richest | 19.8 | 18.6-21.0 |
| **Place of residence** |  |  |
| Urban | 20.9 | 20.0-21.9 |
| Rural | 79.1 | 78.1-80.0 |
| **Region of residence** |  |  |
| Barishal | 5.6 | 5.3-6.0 |
| Chattogram | 22.0 | 21.0-22.9 |
| Dhaka | 23.1 | 22.1-24.1 |
| Khulna | 10.5 | 9.9-11.1 |
| Mymensingh | 7.5 | 6.8-8.1 |
| Rajshahi | 12.6 | 11.9-13.3 |
| Rangpur | 10.8 | 10.2-11.4 |
| Sylhet | 8.0 | 7.4-8.8 |

The distribution of ECDI and its various domains, across several explanatory variables, is presented in Table 2. We observed that nearly 98% of the total children analyzed were on track in the physical domain, followed by 91% in the learning domain, 73% in the socio-emotional domain, and 29% in the literacy-numeracy domain. Overall, approximately 75% of the total children were developmentally on track. Among children who reported participation in the early childhood education program, approximately 86% were developmentally on track, compared to 72% among those who did not participate in the program. We identified an increasing prevalence of overall positive ECDI with the rise in children’s mothers’ education levels and the wealth quintile of the family in which they resided.

**Table 2.** Distribution of early childhood development and its doamins across explanatory variables.

| Exposures | Children who are developmentally track for indicated domains |  |  |  | Child developmentally on track overall, % (95% CI) |
| --- | --- | --- | --- | --- | --- |
|  | Literacy-numeracy domain, % (95% CI) | Physical domain, % (95% CI) | Socio-emotional domain, % (95% CI) | Learning domain, % (95% CI) |  |
| **Total prevalence** | **28.6 (27.7-29.9)** | **98.3 (98.3-98.9)** | **72.8 (71.6-73.9)** | **91.4 (90.8-92.0)** | **74.7 (73.7-75.7)** |
| **Attended in early childhood education** |  |  |  |  |  |
| Yes | 59.6 (56.9-62.2) | 98.9 (98.1-99.3) | 73.9 (71.4-76.1) | 95.3 (94.2-96.3) | 85.8 (83.8-87.6) |
| No | 21.4 (20.3-22.4) | 98.6 (98.2-98.8) | 72.6 (71.4-73.8) | 90.5 (89.7-91.2) | 72.1 (70.9-73.2) |
| **Child's age** |  |  |  |  |  |
| 3 years | 16.3 (15.2-17.5) | 98.4 (97.9-98.7) | 71.0 (69.5-72.5) | 89.7 (88.7-90.6) | 68.7 (67.2-70.1) |
| 4 years | 41.7 (40.0-43.5) | 98.9 (98.5-99.2) | 74.7 (73.3-76.2) | 93.2 (92.4-94.0) | 81.1 (79.8-82.3) |
| **Child's gender** |  |  |  |  |  |
| Male | 27.4 (26.0-28.8) | 98.6 (98.2-98.9) | 69.1 (67.6-70.7) | 91.0 (90.1-91.9) | 71.5 (70.0-72.9) |
| Female | 30.2 (28.7-31.8) | 98.6 (98.2-99.0) | 76.8 (75.4-78.2) | 91.8 (90.9-92.6) | 78.3 (76.9-79.5) |
| **Mother's education** |  |  |  |  |  |
| Pre-primary | 14.7 (12.6-17.0) | 97.8 (96.4-98.6) | 71.9 (69.0-74.6) | 87.6 (85.3-89.6) | 68.3 (65.3-71.1) |
| Primary | 19.5 (17.7-21.4) | 98.2 (97.5-98.8) | 71.5 (69.4-73.6) | 89.5 (88.1-90.8) | 69.3 (67.2-71.5) |
| Secondary | 31.9 (30.3-33.5) | 99.0 (98.7-99.3) | 73.0 (71.4-74.5) | 92.7 (91.8-93.5) | 76.8 (75.4-78.1) |
| Higher | 46.8 (43.7-49.9) | 98.7 (97.8-99.2) | 75.5 (72.7-78.1) | 93.8 (92.1-95.1) | 83.0 (80.6-85.2) |
| **Wealth quintile** |  |  |  |  |  |
| Poorest | 16.7 (15.1-18.5) | 97.8 (96.9-98.4) | 71.7 (69.5-73.8) | 88.5 (86.9-90.0) | 68.1 (65.9-70.3) |
| Second | 23.3 (21.3-25.5) | 98.7 (97.9-99.2) | 70.8 (68.5-72.9) | 90.0 (88.4-91.4) | 71.5 (69.1-73.7) |
| Middle | 30.0 (27.7-32.4) | 98.9 (98.2-99.3) | 72.3 (69.9-74.6) | 91.9 (90.5-93.2) | 75.3 (73.1-77.5) |
| Fourth | 32.5 (30.1-35.0) | 98.5 (97.8-99.0) | 72.1 (69.7-74.5) | 92.8 (91.4-94.0) | 75.7 (73.2-77.9) |
| Richest | 43.0 (40.2-45.9) | 99.4 (98.8-99.7) | 77.4 (74.8-79.8) | 94.3 (92.8-95.4) | 84.0 (81.9-85.9) |
| **Place of residence** |  |  |  |  |  |
| Urban | 34.8 (32.0-37.8) | 98.9 (98.2-99.3) | 74.4 (71.9-76.7) | 92.5 (91.0-93.8) | 78.1 (75.9-80.2) |
| Rural | 27.2 (26.0-28.3) | 98.5 (98.2-98.8) | 72.4 (71.2-73.6) | 91.1 (90.4-91.8) | 73.8 (72.7-75.0) |
| **Region of residence** |  |  |  |  |  |
| Barishal | 30.8 (27.4-34.3) | 99.2 (98.3-99.6) | 64.6 (61.0-68.1) | 88.7 (85.9-90.9) | 68.9 (64.8-70.8) |
| Chattogram | 31.9 (29.4-34.5) | 98.7 (97.9-99.2) | 72.3 (69.8-74.6) | 90.9 (89.5-92.2) | 78.0 (75.9-79.9) |
| Dhaka | 31.9 (29.5-34.4) | 98.8 (98.2-99.3) | 81.8 (79.4-84.0) | 93.6 (92.4-94.6) | 81.6 (79.5-83.6) |
| Khulna | 27.8 (25.1-30.6) | 99.4 (98.9-99.7) | 67.3 (64.7-69.9) | 94.1 (92.6-95.3) | 72.9 (70.3-75.3) |
| Mymensingh | 30.0 (25.5-34.9) | 95.6 (93.0-97.3) | 58.1 (53.6-62.4) | 91.4 (88.8-93.4) | 61.2 (56.6-65.6) |
| Rajshahi | 23.9 (21.2-26.7) | 98.8 (97.8-99.4) | 70.0 (66.7-73.0) | 92.5 (90.5-94.0) | 69.6 (66.3-72.7) |
| Rangpur | 25.9 (23.2-28.8) | 98.2 (97.4-98.8) | 82.8 (8.5-84.8) | 93.4 (91.8-94.8) | 83.7 (51.6-85.6) |
| Sylhet | 21.4 (18.1-25.1) | 99.3 (98.4-99.7) | 66.3 (62.3-70.1) | 80.7 (76.9-84.0) | 61.8 (57.8-65.6) |

**Note:** Presented as row percentages. CI: Confidence intervals.

We identified a 44% (OR, 0.56, 95% CI, 0.47-0.66) lower likelihood of a positive ECDI among children who did not participate in the early childhood education program compared to those who did participate (Table 3). Similarly, the likelihoods of the learning (OR, 0.59, 0.45-0.77) and literacy-numeracy domains (OR, 0.25, 95% CI, 0.21-0.28) of ECDI were lower among children who did not participate in early childhood education compared to those who reported participation in early childhood education.

**Table 3.** Results of multi-level mixed-effect logistic regression models assessing the association of early childhood education with the early childhood development index and its domains among children in Bangladesh, adjusted for covariates.

| Exposures | Domains of Early Childhood Development Index |  |  |  | Early Childhood Development Index |
| --- | --- | --- | --- | --- | --- |
|  | Literacy-numeracy domain | Physical domain | Socio-emotional domain | Learning domain |  |
|  | aOR (95% CI) | aOR (95% CI) | aOR (95% CI) | aOR (95% CI) |  |
| **Attended in early childhood education** |  |  |  |  |  |
| Yes (ref) | 1.00 | 1.00 | 1.00 | 1.00 | 1.00 |
| No | 0.25 (0.21-0.28)*** | 0.97 (0.56-1.68) | 1.05 (0.91-1.22) | 0.59 (0.45-0.77)*** | 0.56 (0.47-0.66)*** |
| **Child's age** |  |  |  |  |  |
| 3 years (ref) | 1.00 | 1.00 | 1.00 | 1.00 | 1.00 |
| 4 years | 3.42 (3.02-3.87)*** | 1.59 (1.60-2.39)** | 1.29 (1.15-1.44)*** | 1.53 (1.28-1.82)*** | 1.96 (1.74-2.19)*** |
| **Child's gender** |  |  |  |  |  |
| Male (ref) | 1.00 | 1.00 | 1.00 | 1.00 | 1.00 |
| Female | 1.15 (1.03-1.28)** | 1.02 (0.70-1.48) | 1.51 (1.36-1.68)*** | 1.09 (0.93-1.29) | 1.48 (1.33-1.64)*** |
| **Mother's education** |  |  |  |  |  |
| Pre-primary (ref) | 1.00 | 1.00 | 1.00 | 1.00 | 1.00 |
| Primary | 1.37 (1.10-1.71)*** | 1.10 (0.64-1.90) | 0.98 (0.82-1.17) | 1.14 (0.89-1.46) | 1.02 (0.85-1.21) |
| Secondary | 2.52 (2.04-3.11)*** | 1.64 (0.94-2.87) | 1.03 (0.87-1.22) | 1.50 (1.17-1.93)*** | 1.35 (1.14-1.60)*** |
| Higher | 4.29 (3.33-5.51)*** | 0.97 (0.47-2.05) | 1.12 (0.90-1.41) | 1.37 (0.96-1.94) | 1.71 (1.35-2.16)*** |
| **Wealth quintile** |  |  |  |  |  |
| Poorest (ref) | 1.00 | 1.00 | 1.00 | 1.00 | 1.00 |
| Second | 1.44 (1.19-1.75)*** | 1.71 (0.98-2.96) | 0.89 (0.76-1.05) | 0.97 (0.77-1.24) | 1.09 (0.92-1.27) |
| Middle | 1.84 (1.52-2.24)*** | 1.80 (0.99-3.29) | 0.96 (0.81-1.14) | 1.24 (0.95-1.61) | 1.23 (1.03-1.45)** |
| Fourth | 1.91 (1.56-2.34)*** | 1.19 (0.65-2.18) | 0.88 (0.74-1.06) | 1.37 (1.03-1.81)** | 1.16 (0.97-1.39) |
| Richest | 2.75 (2.19-3.46)*** | 3.31 (1.37-8.00)*** | 1.22 (0.99-1.51) | 1.84 (1.31-2.58)*** | 1.95 (1.56-2.43)*** |
| **Place of residence** |  |  |  |  |  |
| Urban (ref) | 1.00 | 1.00 | 1.00 | 1.00 | 1.00 |
| Rural | 1.06 (0.89-1.26) | 1.04 (0.57-1.89) | 1.11 (0.94-1.31) | 1.09 (0.84-1.42) | 1.14 (0.96-1.35) |
| **Region of residence** |  |  |  |  |  |
| Barishal (ref) | 1.00 | 1.00 | 1.00 | 1.00 | 1.00 |
| Chattogram | 0.80 (0.60-1.06) | 0.56 (0.18-1.71) | 1.43 (1.10-1.86)*** | 1.09 (0.73-1.61) | 1.51 (1.16-1.97)*** |
| Dhaka | 0.74 (0.55-0.98)** | 0.59 (0.19-1.82) | 2.75 (2.09-3.60)*** | 1.62 (1.08-2.43)** | 2.00 (1.53-2.62)*** |
| Khulna | 0.59 (0.43-0.80)*** | 1.19 (0.31-4.52) | 1.11 (0.84-1.47) | 1.85 (1.17-2.91)*** | 1.16 (0.87-1.53) |
| Mymensingh | 0.93 (0.65-1.31) | 0.18 (0.06-0.55)*** | 0.84 (0.61-1.15) | 1.31 (0.81-2.12) | 0.77 (0.57-1.05) |
| Rajshahi | 0.53 (0.39-0.73)*** | 0.67 (0.20-2.17) | 1.35 (1.03-1.79)** | 1.61 (1.05-2.48)** | 1.07 (0.81-1.41) |
| Sylhet | 0.76 (0.56-1.04) | 0.39 (0.12-1.20) | 2.90 (2.15-3.91)*** | 1.88 (1.21-2.94)*** | 2.68 (1.99-3.61) |
| Rangpur | 0.56 (0.40-0.80)*** | 1.07 (0.27-4.24) | 1.06 (0.78-1.45) | 0.57 (0.37-0.88)** | 0.77 (0.57-1.04) |
| **Model summary** |  |  |  |  |  |
| ICC | 17.82% | 36.92% | 17.45% | 26.33% | 14.50% |
| AIC | 9313.60 | 1310.42 | 10486.16 | 5203.28 | 9832.51 |
| BIC | 9456.58 | 1453.41 | 10629.15 | 5346.27 | 9975.50 |

**Note:** ICC= intraclass correlation; AIC=Akaike's information criterion; BIC=Bayesian information criterion. \*\*\*p<0.01, \*\*p<0.05. aOR: adjusted odds ratio. 95%
CI: 95% Confidence interval. Ref: Reference.

The likelihood of positive ECDI was 1.96 times higher (95% CI, 1.74-2.19) among 4-year-old children compared to their 3-year-old counterparts. Additionally, female children exhibited higher likelihoods of positive ECDI (OR, 1.48, 95% CI, 1.33-1.64) compared to male children. Moreover, children whose mothers had secondary or higher education reported higher likelihoods of positive ECDI than those with mothers having pre-primary education. The likelihood of positive ECDI increased with the wealth quintile of households, reaching statistical significance for children residing in the middle (OR, 1.23, 95% CI, 1.03-1.45) and richest (OR, 1.95, 95% CI, 1.56-2.43) wealth quintiles compared to the poorest quintile. Regionally, compared to children in the Barishal region, those in Chattogram (OR, 1.51, 95% CI, 1.16-1.97) and Dhaka (OR, 2.00, 95% CI, 1.53-2.62) regions exhibited higher likelihoods of positive ECDI. Similar patterns of associations were observed for individual domains of the ECDI.

## Discussion

The objective of this study was to investigate the impact of early childhood education programs on the ECDI and its individual domains. Only 19% of the total children analyzed reported participation in early childhood education. Our analysis revealed that approximately 75% of the total children included in the study reported a positive ECDI, with varying percentages across its domains. Notably, children who did not participate in early childhood education consistently exhibited lower likelihoods of positive ECDI and its independent domains when compared to their counterparts who engaged in early childhood education programs. These findings underscore the crucial role of early childhood education programs in influencing and enhancing various aspects of child development. The results highlight the imperative need for the widespread implementation and accessibility of early childhood education programs to safeguard and promote the enduring well-being of children.

The outcomes of this study indicate that a substantial proportion of children in Bangladesh, approximately 25%, are not on the expected developmental trajectory, amounting to roughly 1 million children when considering the country’s average annual birth rate of 4 million. This estimate is consistent with findings from prior research conducted not only in Bangladesh but also in other LMICs [3, 8, 20]. The magnitude of this significant number highlights a considerable burden on the nation’s resources and underscores the urgent need for targeted interventions. These findings emphasize the critical importance of addressing the challenges associated with poor ECDI outcomes. Past studies consistently link such outcomes to adverse socio-economic and health conditions, encompassing lower parental education levels, disparities in wealth quintiles, instances of malnutrition, and the impact of parental migration [8, 10, 20].

We have found that only one in every five children participates in early childhood education in Bangladesh, despite extensive efforts from both governmental and non-governmental sectors aiming to ensure universal access. The reasons behind this lower participation rate require careful consideration. Potential factors contributing to this phenomenon may include lower awareness among parents about the importance of early childhood education, often associated with lower levels of parental education [15]. Additionally, poverty among parents may contribute to lower participation due to increased engagement in work and a lack of time to focus on their children’s education [21]. Socioeconomic disparities, limited access to educational resources, regional variations in program availability, and cultural influences may also shape parental decisions regarding the enrollment of their children in early childhood education programs [21-23]. Affordability challenges associated with early childhood education, such as the distance from homes, as well as concerns about the quality of early childhood education, also play a role in shaping parental decisions to enroll their children [24, 25]. A comprehensive exploration of these factors is essential to inform targeted strategies that can enhance participation rates and contribute to the fulfillment of the goal of providing early childhood education for all children in the country.

The findings of this study reveal the detrimental impact of non-participation in early childhood education programs on the ECDI. While direct comparisons with existing literature in Bangladesh were not feasible due to its scarcity, the results align with comparable findings developed counties and LMICs [6, 13, 14]. The association between non-participation in early childhood education and adverse ECDI outcomes is complex and influenced by various factors. One possible contributing factor is the potential lack of exposure to structured learning environments and educational stimuli during the critical early years of development [26]. Early childhood education programs often provide a foundation for cognitive, social, and emotional development, and the absence of such experiences may hinder overall developmental progress [27, 28]. Moreover, disparities in socio-economic conditions, parental education levels, and access to resources could play a role [28]. Children who do not participate in early childhood education programs may face additional challenges related to family income, educational support at home, and overall household well-being [21, 29-31]. Addressing these socio-economic determinants becomes crucial in understanding and mitigating the negative effects observed in ECDI outcomes. Furthermore, cultural and contextual factors might contribute to the observed associations [30]. Differences in cultural attitudes towards education and the availability of early childhood education opportunities could influence participation rates and subsequently impact developmental outcomes [32].

The policy implications derived from this study underscore the imperative to fortify early childhood education programs in Bangladesh. This necessitates an enhancement of accessibility by ensuring the availability of early childhood education facilities near healthcare facilities and an overarching improvement in the quality of these programs. A strategic focus on raising parental awareness regarding the significance of early childhood education is paramount to overcoming barriers to participation. Concurrently, addressing health issues among children, particularly malnutrition, should be a focal point. Efforts should be directed towards creating awareness among parents about the crucial link between their children’s health and their engagement in early childhood education. This comprehensive approach, encompassing strengthened program accessibility, heightened quality, increased parental awareness, and targeted health interventions, is crucial for maximizing the impact of early childhood education initiatives in Bangladesh.

This study exhibits both strengths and limitations that warrant consideration. The data employed in this research is derived from a cross-sectional survey, indicating that the findings are correlational rather than establishing causation. Furthermore, the reliance on self-reported data in the surveyed information introduces the possibility of recall bias. Additionally, while this study adjusts for various factors influencing the relationship between early childhood education and the ECDI, other crucial factors such as the availability and accessibility of early childhood education programs might contribute to this association, emphasizing the necessity of their inclusion in future research. Despite these limitations, this study marks the first attempt in Bangladesh to explore the association between early childhood education and ECDI. Leveraging a large-scale, nationally representative dataset with comprehensive covariate adjustments at both the child and parental levels enhances the robustness of our findings. As such, the outcomes of this study hold significant implications for the development of national-level policies and programs.

## Conclusion

One in every five children in Bangladesh reported participating in early childhood education. Non-participation in early childhood education programs was found to be associated with poor ECDI outcomes. These findings underscore the imperative of ensuring universal access to early childhood education for all children. It is crucial to implement policies and programs aimed at achieving this goal. To achieve widespread access, early childhood education facilities need to be strategically located near parental homes, and a simultaneous focus on enhancing program quality is essential. Equally important is the need to elevate parental awareness regarding the significance of early childhood education. A comprehensive approach, encompassing strategic program placement, quality enhancement, and heightened awareness initiatives, is essential to bridge the current gap in early childhood education participation and pave the way for improved child development outcomes in Bangladesh.

## Data Availability

The authors have made the data associated with this study accessible to interested researchers. To access the raw data file, interested researchers need to submit a research proposal to UNICEF through the following link: https://mics.unicef.org/surveys.

## Declarations

### Ethics Approval

The surveys we analysed were conducted by UNICEF as part of their efforts to gather evidence on various aspects of children’s and maternal health. Before initiating these surveys, ethical approval was obtained from the relevant countries’ ethical approval boards, as well as UNICEF’s own board. Informed written consent was obtained all respondents before survey is conducted. Children’s data were collected from their parents or actual care givers. The data, provided to us in a non-identifiable form, was shared in response to our expression of interest in conducting this research, which involved submitting a research proposal. Others interested in accessing this data can follow a similar procedure. Since we only received non-identifiable data, no further ethical approval was required.

### Consent for Publication

Not applicable.

### Authors’ Contribution

Khanam SJ and Khan MN developed the study concept. Alam MB and Khanam SJ analyzed the data and wrote the first draft of this manuscript. Khan MN critically revised the manuscript. All authors approved the final version of the manuscript.

## Acknowledgments

The authors express their gratitude for the assistance provided by the Department of Population Science at Jatiya Kabi Kazi Nazrul Islam University, Bangladesh, where this study was undertaken.

